## Supplementary figures and images for "RISKS OF NEONATAL MORTALITY IN NEONATES WITH NEONATAL ENCEPHALOPATHY IN A TERTIARY NEWBORN CARE UNIT IN ZIMBABWE OVER A 12-MONTH PERIOD"

### Supplemental Material 1

Supplementary Material 1:Data flowchart

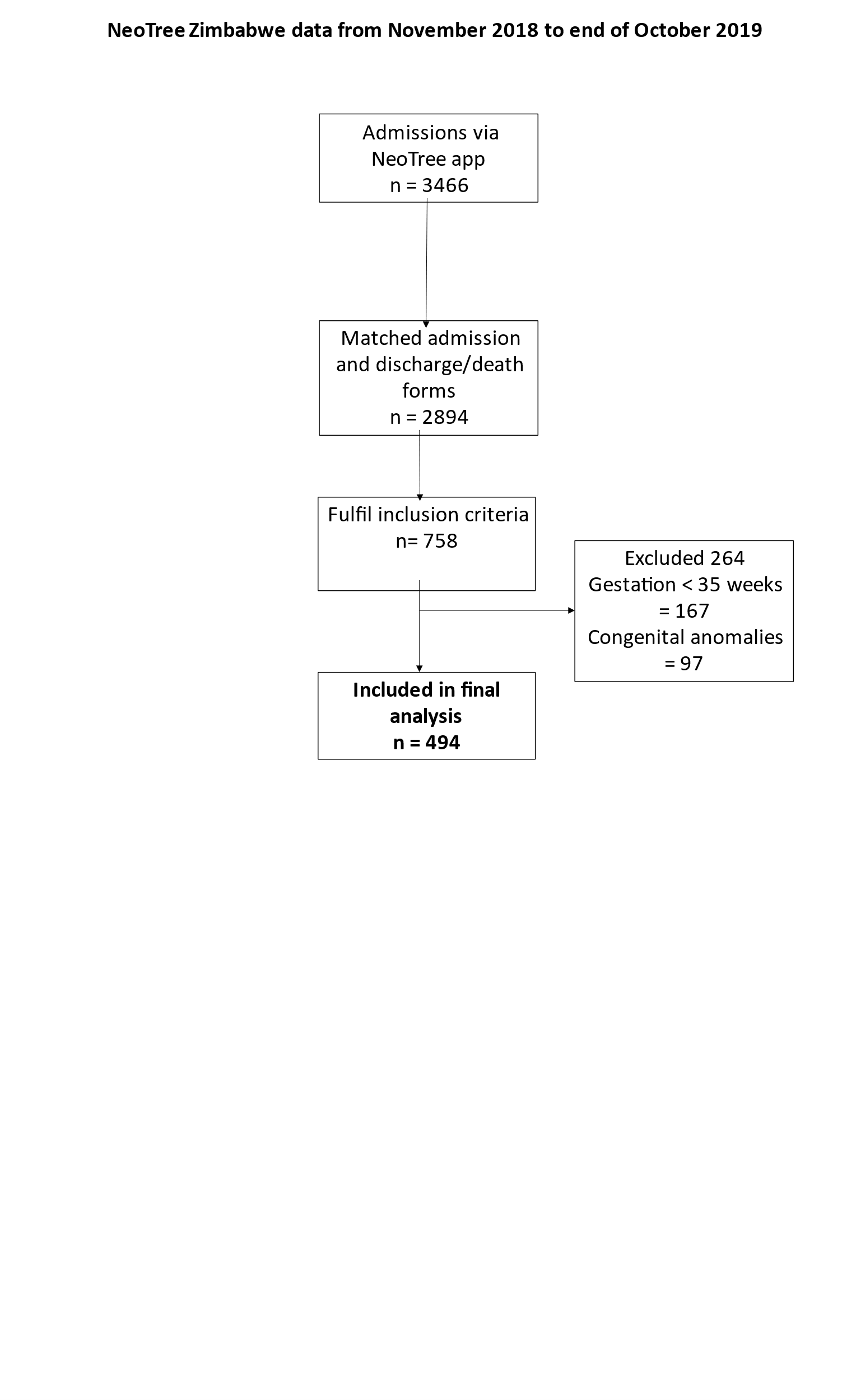
